# Evaluating GPT-4o Model Proficiency and Clinical Reasoning for Antimicrobial Stewardship in Dentistry

**DOI:** 10.64898/2026.09.01.26361980

**Authors:** Madison Dick, Sreenath Madathil, Aditya Patel, Harsimran Singh Kapoor, Mridul Sharma, Zaneta D’Souza, Shahul Hameed, Mohammad Abu-Samak, Amin Najirad, Dirgham Dwairi, Osama Radaideh, Belinda Nicolau

## Abstract

**Objectives:** Dentists prescribe approximately one in ten antibiotics worldwide, yet antimicrobial stewardship (AMS) remains underemphasized in dental education. Large language models (LLMs) may support AMS training, but their proficiency and clinical reasoning in this context remain unclear. We evaluated GPT-4o’s accuracy and clinical reasoning on dental antibiotic prescribing questions, stratified by question difficulty.

**Methods:** We assembled 125 multiple-choice questions on dental antibiotic prescribing from eight peer-reviewed studies (2017–2023). GPT-4o answered each question and generated a clinical justification. Accuracy was assessed against source-study answer keys and examined across difficulty quartiles. Justifications were evaluated using an adapted 12-axis human-evaluation framework assessing scientific consensus, extent and likelihood of harm, inappropriate and missing content, bias, and both correct and incorrect comprehension, retrieval, and reasoning. Prophylaxis-specific questions were analysed separately.

**Results:** GPT-4o correctly answered 72% of questions. Accuracy remained relatively stable across difficulty quartiles (78%, 78%, 65%, 70%). Experts rated 95.4% of justifications positively across the 12 axes. Comprehension, retrieval, and reasoning each exceeded 96.2% positive ratings. Missing content was the main weakness (7.8%), and 7.1% of justifications showed a moderate-to-severe potential for harm. Performance on prophylaxis-specific questions (98.1%) exceeded non-prophylaxis questions (93.0%).

**Conclusions:** GPT-4o demonstrated moderate-to-high proficiency and clinically defensible reasoning in dental antibiotic prescribing questions. However, residual risks indicate that it is not suitable for unsupervised clinical use but shows potential as a supervised AMS educational tool.

## Introduction

Antimicrobial resistance (AMR) is one of the most consequential public health challenges of the twenty-first century. Bacterial AMR was directly responsible for an estimated 1.27 million deaths and associated with 4.95 million deaths globally in 2019, exceeding the combined annual mortality from HIV and malaria [1]. Projections estimate 1.91 million annual AMR-attributable deaths by 2050 and 39.1 million cumulative deaths between 2025 and 2050 [2], with cumulative economic costs reaching US$100 trillion if current trajectories continue [3]. The 2024 World Health Organization (WHO) Bacterial Priority Pathogens List reaffirmed AMR as one of the top ten global health threat requiring concerted action across all prescribing disciplines [4].

Dentistry, as a major prescribing discipline, is a substantial contributor to global antibiotic consumption. Dentists are responsible for approximately 10% of all antibiotic prescriptions worldwide [5]. National data show similar patterns, with dental prescribing accounting for 9.8%– 12.1% of outpatient antibiotic prescriptions in the United States [6] and 10.8% of primary-care antibiotic prescriptions in England [7]. While antibiotics are necessary in certain clinical situations, a large proportion of dental prescribing has been deemed inappropriate. In the United States, more than 80% of antibiotics prescribed for dental procedure prophylaxis between 2011 and 2015 were judged unnecessary [8]. A recent systematic review highlighted that up to 90% of dental antibiotic prescriptions may be inappropriate [9]. Furthermore, during the COVID-19 pandemic, dental antibiotic prescribing increased by 25% as access to in-person care was deferred toward remote prescribing [10].

These prescribing failures persist despite strong evidence that systemic antibiotics offer no added benefit over operative intervention alone [11]. Clinical guidelines support this evidence and provide recommendations for the management of pulpal and periapical infection [12], prosthetic-joint prophylaxis [13], infective endocarditis prophylaxis [14], and endodontic antibiotic use [15]. The FDI World Dental Federation has also identified the dental team as essential to AMR mitigation [16]. Despite this, surveys of dental practitioners and students consistently report gaps in guideline knowledge and low confidence in prescribing decisions, particularly for antibiotic prophylaxis [17-21]. These findings indicate that inappropriate prescribing reflects not only knowledge gaps but also limitations in training and clinical decision making.

Large language models (LLMs) have emerged as a potential tool for medical education and clinical decision support. Recent studies show that these models can perform well on standardized assessments. For example, GPT-4 exceeded the United States Medical Licensing Examination pass threshold by more than 20 percentage points [22], and Med-PaLM 2 reached 86.5% on MedQA, with physicians preferring its responses on most clinical-utility axes [23]. Similar patterns emerge in dentistry, with GPT-4 having passed both the United States Integrated National Board Dental Examination and the United Kingdom Overseas Registration Examination, scoring 72.1% on a 1,461-question composite test [24]. GPT-4o, the model evaluated in the present study, has subsequently outperformed GPT-4 on the Japanese National Dental Examination (73.8% vs 63.3%) [25] and matched the pass mark of human candidates on the Polish Final Dentistry Examination for factual questions, although it fell short on case-based reasoning [26]. This trajectory, while potentially transformative for medical and dental education, also highlights persistent risks of hallucination, bias, and missing context [27, 28].

Two limitations of this evidence base motivate the present study. First, dental licensing exams sample broadly across clinical and basic sciences and do not assess the prescribing judgements that drive AMR in practice. Second, the only published evaluation of an LLM on dental antibiotic decision-making to date examined 28 true/false questions derived from the 2021 American Heart Association (AHA) infective endocarditis guidance and found that several models relied on outdated recommendations [29]. Evidence from outside dentistry have raised additional safety concerns. In a prospective bloodstream infection cohort, only 36% of GPT-4 antibiotic recommendations were judged optimal and 5% were considered potentially harmful [30]. A recent review also concluded that LLMs are not yet ready for autonomous antimicrobial prescribing [31]. Therefore, it remains unclear whether contemporary LLMs can provide accurate and clinically defensible recommendations across dental antibiotic prescribing decisions, and whether they could be safely used as supervised antimicrobial stewardship (AMS) educational tools.

We therefore evaluated GPT-4o on 125 multiple-choice questions drawn from eight peer-reviewed studies of dentists’ antibiotic prescribing knowledge. We assessed GPT-4o accuracy stratified by item difficulty (defined by respondent accuracy in the source studies as an external proxy) and evaluated its written justifications using a 12-axis expert framework adapted from Singhal et al. 2023 [32]. We hypothesised that GPT-4o would achieve moderate-to-high accuracy on dental antibiotic prescribing questions, maintain accuracy across the difficulty gradient, and show identifiable areas of weakness in clinical reasoning that could inform targeted educational use.

## Materials and Methods

### Question Dataset

We conducted a systematic search of literature to identify peer-reviewed studies that had administered multiple-choice questionnaires on antibiotic prescribing to dentists or dental students. The search was conducted across the databases PubMed, OVID Medline, Cochrane, Scopus, and Web of Science. Forward and backward citation searching was additionally employed. Search terms centered on the following key concepts: (i) antibiotic prescribing; (ii) dentistry; and (iii) survey. Inclusion criteria required studies to be original surveys of dentists or dental students with published multiple-choice questions, including explicit reporting of the answer key or expert-consensus correct response. Eight studies meeting these criteria, published between 2017 and 2023, contributed a combined 125 questions (Table 1) [17-19, 33-37]. Two dental clinicians (SH, ZD) with AMS expertise independently screened each question for content validity, clinical relevance, and unambiguous wording; disagreements were resolved by consensus. Per-question respondent accuracy reported in each source study served as a proxy for question difficulty.

**Table 1.** Source studies and contributed multiple-choice questions (n = 125).

| Source study | Year | Region / Setting | Questions, n |
| --- | --- | --- | --- |
| Al-Johani et al. [33] | 2017 | Saudi Arabia (general dentists, Jeddah) | 5 |
| Mansour et al. [17] | 2018 | Lebanon (general dentists) | 36 |
| Teoh et al. [18] | 2019 | Australia (general dentists) | 11 |
| Baudet et al. [19] | 2020 | France (nationwide dentists) | 17 |
| Šimundić Munitić et al. [34] | 2021 | Croatia (endodontists / dentists) | 9 |
| Khalil et al. [35] | 2022 | Sweden (emergency dental care, Stockholm) | 3 |
| Ealla et al. [36] | 2023 | India (general dentists, Hyderabad) | 14 |
| Sirinoglu Capan et al. [37] | 2023 | Türkiye (paediatric dentistry) | 30 |
| **Total** |  |  | **125** |

### LLM Response Generation

We evaluated GPT-4o (OpenAI; model snapshot accessed July 2024) via the OpenAI Application Programming Interface (API) using a single, uniform prompt template. GPT-4o was selected on three grounds. First, GPT-4o is among the most widely accessible general-purpose large language models, available to health-professions students and clinicians through both consumer and developer interfaces, making it an ecologically valid target for an evaluation framed around antimicrobial stewardship education. Second, GPT-4o is freely accessible through both consumer and developer interfaces, with no institutional licensing barriers to adoption by dental learners or faculty. Third, at the time of evaluation GPT-4o was among the leading publicly available general-purpose models on standardised medical and dental knowledge benchmarks [24-26], providing a representative estimate of current default-configuration LLM performance. For every question, the model received the verbatim question stem followed by all answer options and was instructed to (i) select a single best option and (ii) provide a paragraph-length justification for its choice. Default model parameters were used, and no retrieval augmentation, system prompt, or chain-of-thought scaffolding was applied, to characterise default-configuration performance as encountered by the typical educational user. Each question was queried in an independent API call to prevent context leakage between items.

### Accuracy Scoring and Difficulty Stratification

Each GPT-4o answer was scored binarily (1 = correct, 0 = incorrect) against the answer key from the source study; overall accuracy was calculated as the proportion of correct responses. To characterise GPT-4o performance as a function of question difficulty, questions were ranked by respondent accuracy reported in each source study and divided into four quartiles of approximately equal size (n ≈ 31 per quartile). Quartile 1 contained the questions respondents in the source studies answered most accurately; Quartile 4 contained the most difficult questions. Respondents in the source studies were heterogeneous, comprising general dentists, specialists, and dental students across different countries and time periods, and respondent accuracy was used solely as an external proxy for question difficulty rather than as a directly comparable performance group. GPT-4o accuracy was then computed within each quartile to examine whether its performance varied with item difficulty.

### Expert Evaluation of Clinical Justifications

Justifications were evaluated by six dental experts (ZD, SH, MAS, AN, DD, OR) using a 12-axis human-evaluation instrument adapted from Singhal et al. (2023) [32]. The 125 questions were split into two non-overlapping sets to control rater fatigue, and each set was independently rated by a separate panel. The instrument captured 12 axes: (1) agreement with scientific and clinical consensus, (2) extent of potential harm, (3) likelihood of harm, (4) presence of inappropriate content, (5) presence of missing content, (6) possibility of bias, (7) correct comprehension, (8) incorrect comprehension; (9) correct retrieval, (10) incorrect retrieval, (11) correct reasoning, and (12) incorrect reasoning. A justification was scored as having a positive expert assessment on a given axis when raters reached majority agreement on the favourable response option (e.g., “no harm” as opposed to “death or severe harm” or “moderate or mild harm”). A representative evaluation prompt is provided in Figure 1.

**Figure 1.**
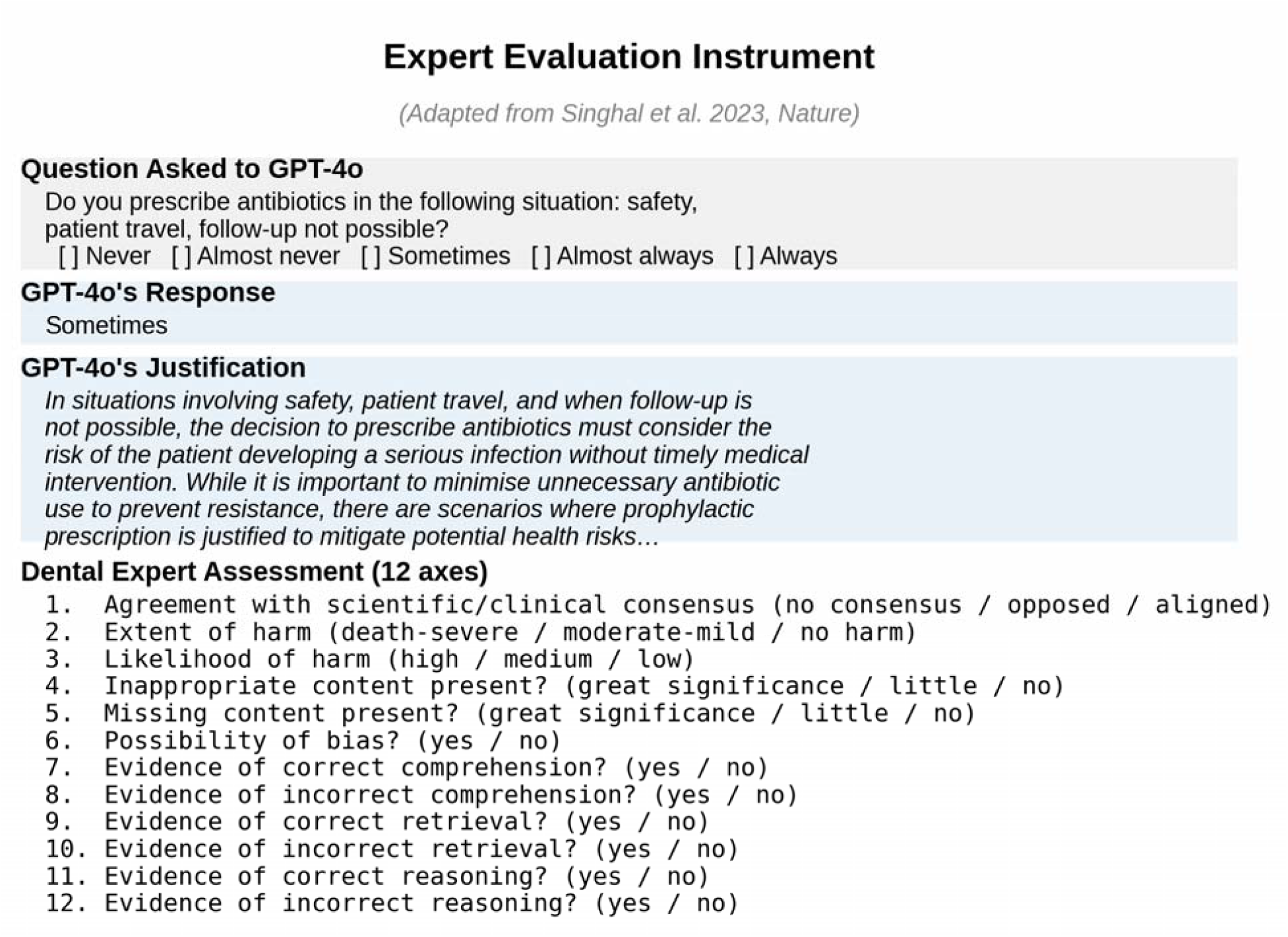
Representative expert-evaluation prompt. ^*^Each GPT-4o response was rated by dental-content experts on 12 axes adapted from Singhal et al. (2023): scientific consensus, extent of harm, likelihood of harm, inappropriate content, missing content, possibility of bias, and the correctness of comprehension, retrieval, and reasoning (each scored separately as correct and incorrect).

### Analysis of Clinical Justifications

Per-axis positive-rating proportions from expert assessment were computed across the 125 questions and 12 axes (1,500 axis-question combinations). Questions that received negative ratings from a majority of evaluation axes were identified as potential areas of weakness. Questions relating to antibiotic prophylaxis were tagged a priori during dataset construction and analysed as a separate stratum, motivated by the well-documented disproportionate share of dental prescribing errors attributable to prophylaxis [8, 20]. Prophylaxis status was assigned based on the clinical content and wording of each question, with questions explicitly addressing whether antibiotic prophylaxis should be prescribed classified as prophylaxis questions.

### Reporting and Ethics

This study evaluated an artificial-intelligence system using already-published, anonymized multiple-choice questions rated by faculty raters in their educational capacity and was determined not to constitute human-subjects research by the McGill University Faculty of Medicine and Health Sciences Research Ethics Office. Reporting follows the Transparent Reporting of a multivariable prediction model for Individual Prognosis Or Diagnosis (TRIPOD) – LLM extension guidelines [38].

## Results

### Overall Accuracy

GPT-4o correctly answered 90 of 125 multiple-choice questions, for an overall accuracy of 72.0%, indicating moderate-to-high proficiency on the specific domain of antibiotic prescribing.

### Difficulty-Stratified Performance

Across the four difficulty quartiles, GPT-4o achieved 78% accuracy in Quartile 1 (easiest questions), 78% in Quartile 2, 65% in Quartile 3, and 70% in Quartile 4 (most difficult questions). Accuracy was relatively stable across the difficulty gradient, with no monotonic decline as questions became more difficult for the respondents in the original studies. Notably, GPT-4o maintained 70% accuracy in Quartile 4, the quartile of questions answered correctly by the smallest proportion of respondents in the source studies suggesting that its performance did not collapse on the items that human respondents found most challenging.

### Quality of Clinical Justifications

Across the 12 evaluation axes and 125 questions, GPT-4o’s clinical justifications received positive expert ratings on 95.4% of axis-question combinations. Per-axis rates ranged from 92.9% to 98.9% (Table 2). The strongest axes were correct comprehension, correct retrieval, and correct reasoning, each above 96.2% positive ratings. The weakest axes were missing content, scientific consensus, and extent of harm. On missing content, 5.7% of justifications were judged to omit information of little clinical significance and 2.1% to omit information of great clinical significance. On consensus, 4.6% of justifications were judged opposed to clinical consensus, and on harm, 7.1% of justifications carried a potential for moderate or severe harm if acted upon without supervision. Possibility of bias was identified in fewer than 5% of justifications, predominantly in questions involving paediatric dosing or penicillin allergy.

**Table 2.**
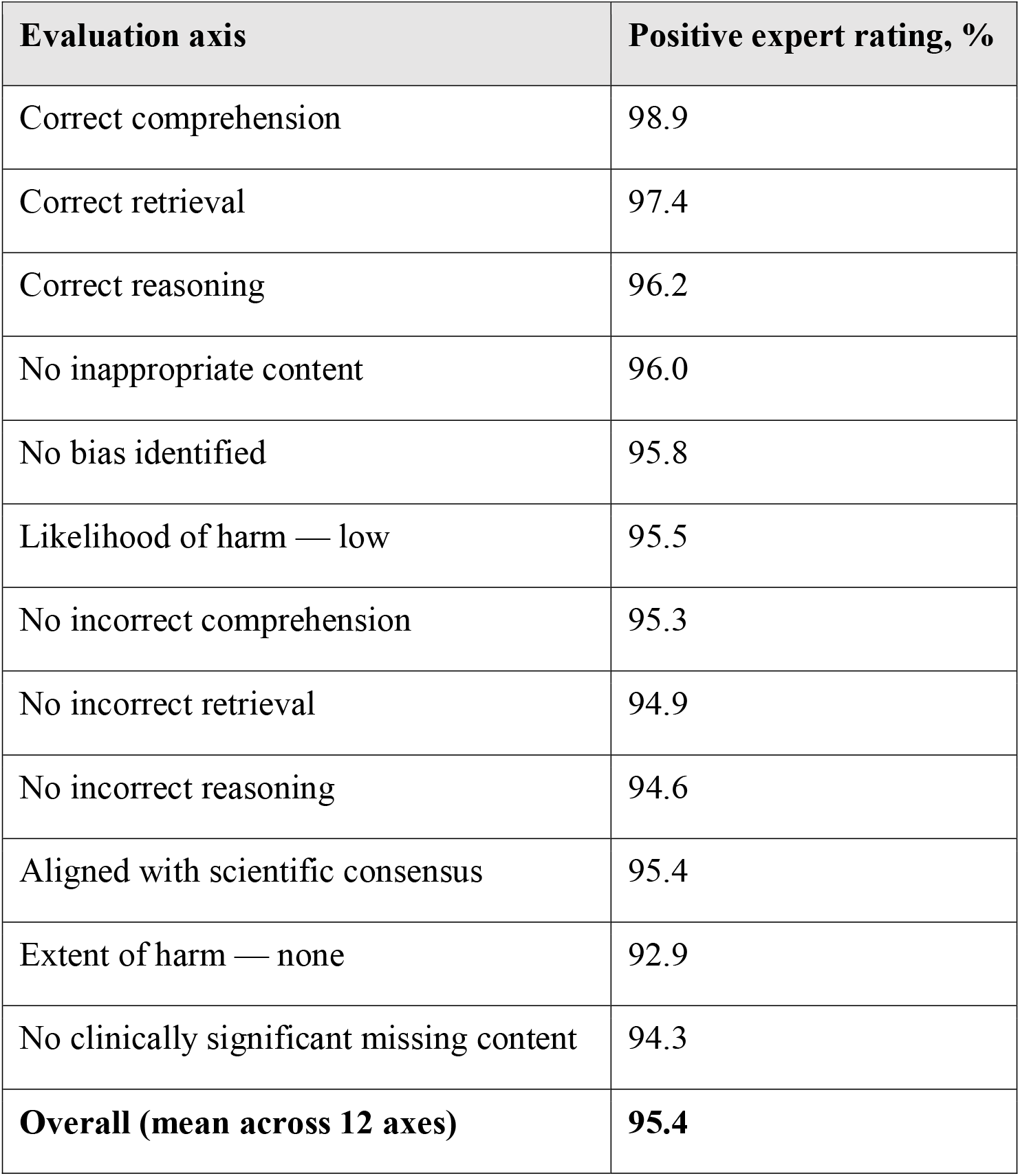
Expert evaluation of GPT-4o clinical justifications across 12 axes (n = 125 questions × 12 axes = 1,500 axis-question combinations).

### Topic-Specific Performance

Questions focusing on antibiotic prophylaxis received positive expert ratings on 98.1% of axis-question combinations, compared with 93.0% for non-prophylaxis questions, a 5.1-percentage-point absolute advantage for prophylaxis. The clinical-content categories on which a majority of evaluation axes registered expert disagreement were: indications for antibiotic adjuncts in routine restorative care; necrotic pulp with acute apical abscess; acute necrotizing ulcerative gingivitis; osteitis; salivary-gland infection; tooth avulsion; cellulitis; periodontal regeneration and pre-implantation surgical infection; fever in combination with deep carious lesions; endodontic infection in patients with documented penicillin allergy; and the heterogeneous “safety, patient travel, and follow-up not possible” cluster. These categories did not cluster within a single anatomical site or single source study, indicating dispersed rather than systematic gaps in GPT-4o’s dental antimicrobial knowledge.

## Discussion

This study evaluates GPT-4o across 125 dental antibiotic prescribing questions drawn from eight published instruments; previous evaluations in this domain have been narrower in scope, the closest comparator having assessed 28 true/false questions on infective endocarditis prophylaxis [30]. Three findings are central. First, GPT-4o achieved 72.0% accuracy across 125 questions spanning eight published instruments, a level comparable to reported GPT-4o performance on whole-curriculum dental licensing examinations [25] but obtained on the specific judgements that drive AMR in dental practice. Second, GPT-4o accuracy was stable across difficulty quartiles, including the quartile of items that source-study respondents found most challenging; this flat difficulty profile indicates that GPT-4o does not collapse on the items where prescribing knowledge has previously been shown to be weakest, and identifies the AMS-relevant scenarios where supervised decision support could be most valuable. Third, expert raters judged 95.4% of GPT-4o’s clinical justifications as defensible across a 12-axis adaptation of the Singhal et al. [32] evaluation framework, with comprehension, retrieval, and reasoning all exceeding 96.2% positive ratings. Performance on antibiotic prophylaxis, the single category most associated with dental over-prescribing [8, 20] was the strongest of all topic strata at 98.1%.

These findings situate GPT-4o between two markers in the recent literature. On the optimistic side, Med-PaLM 2 reached 86.5% on MedQA and was preferred by physicians over peer answers on 8 of 9 clinical axes [23]; GPT-4o has matched or exceeded GPT-4 on multiple dental licensing benchmarks [24, 25]; and the per-axis ratings reported here approach the standards expected of teaching materials. On the cautionary side, Maillard et al. [30] found that only 36% of GPT-4 antibiotic recommendations for bloodstream infection were optimal and 5% were potentially harmful; Rewthamrongsris et al. [29] showed that several LLMs cited outdated infective endocarditis guidance; and Giacobbe et al. [31] concluded that LLMs are not yet ready for autonomous antimicrobial prescribing. Our results are consistent with both signals: GPT-4o is materially more competent than the autonomous-prescribing literature alone would suggest, but a 7.1% rate of justifications carrying moderate-to-severe harm potential, together with a 2.1% rate of clinically significant missing content, places it firmly outside the range of safe unsupervised use.

The pattern of dispersed weakness deserves comment. The clinical scenarios on which GPT-4o’s justifications drew majority expert disagreement acute necrotizing ulcerative gingivitis, salivary-gland infection, osteitis, avulsion, cellulitis, peri-implant and periodontal regeneration infection, and complex modifiers such as documented penicillin allergy or impossible follow-up share three properties: they are clinically uncommon relative to the bulk of dental practice, they require integration across multiple guideline domains, and they often turn on patient-specific risk modifiers rather than the underlying infection alone. This pattern is consistent with the failure mode reported by Jaworski et al. [26], in which GPT-4o scored 72.87% on factual dental questions but only 36.36% on case-based reasoning. The implication is that LLMs trained on broad text corpora reproduce population-level guideline knowledge with high fidelity but generalise less reliably to the long tail of patient-specific decisions where dental AMR errors most commonly arise.

Three educational implications follow. First, GPT-4o is a credible candidate as a supervised question-bank authoring tool and as a formative-assessment tutor for difficult questions, where the consequences of error are buffered by the supervisory loop. Second, deployments must mitigate the missing-content and harm signals through structured retrieval against authoritative sources for example, the American Dental Association, American Association of Endodontists, AHA, FDI World Dental Federation, and Scottish Dental Clinical Effectiveness Programme guidelines [12-16] rather than reliance on default-configuration model output. Third, evaluation pipelines must move beyond simple accuracy. The 12-axis Singhal framework applied here, augmented with prophylaxis stratification and difficulty-quartile analysis, is the minimum granularity needed to distinguish a model that is competent on average from one that is safe in deployment. We endorse the TRIPOD-LLM reporting standard [38] and the WHO ethics-and-governance framework for large multimodal models in health [39] as the appropriate baseline for this work.

Our study has several limitations. The questions were drawn from existing instruments and therefore over-represent the topics those instruments emphasised, particularly prophylaxis and paediatric prescribing; this constrains generalisation to underrepresented domains such as periodontal-abscess management and oncology-related prescribing. Respondent accuracy used to define difficulty quartiles was extracted from heterogeneous source studies (general dentists, specialists, and dental students across multiple countries and time periods) rather than measured prospectively in a single cohort; it serves as an external proxy for item difficulty rather than as a directly comparable performance group. Expert raters drew from a single institution; survey fatigue may have attenuated qualitative engagement on later questions, even with the two-set design. We evaluated a single model snapshot at a single timepoint, with no retrieval augmentation, no system prompt, and no chain-of-thought scaffolding; performance under richer configurations is likely higher but was not the question of interest. Finally, the present study evaluates performance on multiple-choice and free-text justification, not patient-facing clinical interaction; safety in real prescribing workflows requires a dedicated trial.

## Conclusion

GPT-4o demonstrated moderate-to-high accuracy on 125 antibiotic-prescribing questions and maintained accuracy across difficulty quartiles, including the quartile of items that source-study respondents found most challenging. Expert raters judged 95.4% of its written clinical justifications as defensible across a 12-axis evaluation framework, with antibiotic prophylaxis, the single largest source of dental antibiotic over-prescribing emerging as the model’s strongest category. Residual signals of harm (7.1%) and clinically significant missing content (2.1%) preclude unsupervised clinical use but are compatible with the supervised, education-first deployment pattern that the dental AMS literature identifies as the priority. Future work should evaluate retrieval-augmented and guideline-grounded configurations against guideline-keyed reference standards, extend evaluation to patient-facing decision support under prospective conditions, and characterise model drift as new model versions are released.

## Data Availability

All data generated and analysed in the current study are available from the corresponding author upon reasonable request.

